# Effect of public health interventions during the first epidemic wave of COVID-19 in Cyprus: a modelling study

**DOI:** 10.1101/2021.01.02.20248980

**Authors:** Ilias Gountas, Annalisa Quattrocchi, Ioannis Mamais, Constantinos Tsioutis, Eirini Christaki, Konstantinos Fokianos, Georgios Nikolopoulos

## Abstract

**Background:** Cyprus addressed the first wave of SARS CoV-2 (COVID-19) by implementing non-pharmaceutical interventions. The aims of this study were: a) to estimate epidemiological parameters of this wave including infection attack ratio, infection fatality ratio, and case ascertainment ratio, b) to assess the impact of public health interventions, and c) to examine what would have happened if those interventions had not been implemented.

**Methods:** A dynamic, stochastic, individual-based Susceptible-Exposed-Infected-Recovered (SEIR) model was developed to simulate COVID-19 transmission and progression in the population of the Republic of Cyprus. The model was fitted to the observed trends in COVID-19 deaths and intensive care unit (ICU) bed use.

**Results:** By May 8 2020^th^, the infection attack ratio was 0.31% (95% Credible Interval (CrI): 0.15%, 0.54%), the infection fatality ratio was 0.71% (95% CrI: 0.44%, 1.61%), and the case ascertainment ratio was 33.2% (95% CrI: 19.7%, 68.7%). If Cyprus had not implemented any public health measure, the healthcare system would have been overwhelmed by April 14^th^. The interventions averted 715 (95% CrI: 339, 1235) deaths. If Cyprus had only increased ICU beds, without any social distancing measure, the healthcare system would have been overwhelmed by April 19^th^.

**Conclusions:** The decision of the Cypriot authorities to launch early non-pharmaceutical interventions limited the burden of the first wave of COVID-19. The findings of these analyses could help address the next waves of COVID-19 in Cyprus and other similar settings.

## Introduction

On January 30^th^ 2020, the World Health Organization declared the outbreak of 2019 Coronavirus disease (COVID-19) a Public Health Emergency of International Concern (1), and on March 11^th^, a pandemic. The threat of COVID-19 to global health comes from the high percentage of the population that is not immune to the new pathogen (SARS-CoV-2), the high transmissibility of SARS-CoV-2 in comparison to other respiratory viruses, and the potential substantial burden on healthcare systems (2, 3). In the absence of effective therapeutics or vaccines, the majority of countries responded to COVID-19 with non-pharmaceutical interventions (NPI) including social distancing measures, in order to limit the viral spread, to ensure that the healthcare system would not be overwhelmed, and to prevent mass casualties.

The first COVID-19 cases in the Republic of Cyprus (∼876000 residents in the government-controlled area) were detected on March 9^th^, 2020 followed by a surge in diagnoses that peaked in late March to early April. The response to the outbreak included several timely social distancing measures. More specifically, on March 10^th^ schools and universities closed and, five days later, the access to the country was restricted and entertainment areas closed. On March 24^th^, most retail services closed and on March 31^st^ social gatherings were prohibited (4). A key public health response was also the wide test-trace-isolate strategy. During the first epidemic wave (by May 8^th^ 2020), the testing rate in the Republic of Cyprus was 8932 per 100,000 population, which was significantly higher than in other European countries (e.g., the testing rate was 3018, 1217 and 926 per 100,000 population for the United Kingdom, the Netherlands, and Greece, respectively over the same period (4). Finally, in order to meet the healthcare challenges anticipated by COVID-19, the number of COVID-19-designated intensive care unit (ICU) beds increased from 27 at the beginning of the outbreak to more than 50 by May 8^th^ (5). By May 8^th^, only 20 COVID-19-related deaths (2.28 deaths per 100,000 population) were reported (4, 5).

Although the country’s response seems successful in controlling the first wave of COVID-19, it is important to quantitatively assess the effectiveness of the interventions in the Republic of Cyprus. To this end, mathematical models can provide important insights (3, 6-10). In this study, we report the results of a dynamic COVID-19 modeling approach. Our aims were: a) to provide epidemiological estimates for the first wave of COVID-19 in the Republic of Cyprus, b) to estimate the impact of NPI, and c) to examine possible outcomes in the hypothetical scenario where these measures were not implemented.

## Methods

### Description of the mathematical model

To simulate SARS-CoV-2 transmission and progression, a discrete time, stochastic, individual-based model, which categorizes the population into susceptible, exposed, infectious, or recovered (SEIR) individuals, was developed in programming language C++ (Dev-C++ v.5.11) (Supplementary, page 2, Figure S1).

Every day, susceptible individuals might contact infected people and enter the exposed disease state before they become infectious. Infectiousness may start at least 1 to 2 days before the onset of symptoms (2, 11, 12). Asymptomatic patients or patients with mild symptoms could recover from the disease after 5-7 days (8, 13). Patients with severe or critical disease may need to enter a healthcare facility.

According to COVID-19 surveillance data in the Republic of Cyprus from March 1^st^ to May 8^th^,2020, the median time from symptom onset to hospital admission of patients with severe symptoms (i.e., requiring hospitalization) was 6.5 days (Interquartile Range (IQR): 4.5-9.5 days) (Table 1). The median time from symptom onset to hospital admission for patients who were later admitted to ICU was 5.5 days (IQR: 3.5-10 days). The overall median length of stay in ICU was 13.5 days (IQR: 8-28 days) (4, 5). Cases in the ICU would either die or recover after spending a period of recovery time in a hospital bed. The probability of dying in the ICU bed during the first wave was estimated at 41% (4, 14). Further details about the description of the model and the calibration procedure are available in the supplementary material. All healthcare inputs of the model were retrieved from publicly available data (4).

**Table 1:** Model parameters, assumptions and references

| Parameters | Value | References |
| --- | --- | --- |
| Cyprus population | 876000 | Statistical Service of Cyprus (CYSTAT) Demographic Statistics 2018; Ministry of Finance: Cyprus, 2019 |
| Incubation period | 5 days | (15) |
| Latent period<sup>†</sup> | 4 days (e.g., 1 day prior to the occurrence of symptom) | (11) |
| Duration of infectious period until recovery | 6 days | (2) |
| Duration from the development of symptoms to recovery | 5 days | (2) |
| Median length of stay in Intensive Care Unit (ICU) bed | 13.5 days | (4) |
| Disease mortality in ICU | 41% | (4, 14) |
| Proportion of infections that will be either asymptomatic or mild ($F_m$)<br>Proportion of infections that will progress to severe disease ($F_s$)<br>Proportion of infections that will progress to critical disease ($F_c$) | $F_m = 86.0\%$<br>$F_s = 11.5\%$<br>$F_c = 2.5\%$ | Fit to the observed deaths, hospitalized cases, and use of ICU beds |
| Duration of hospitalization | 15 days | (4) |
| Duration of ICU and hospitalization | 20 days |  |

### Model parameterization and examined scenarios

The validity of estimates from modeling studies depends on the quality of the data to which models are calibrated (3, 9, 16). Under-ascertainment of COVID-19 cases has been recognized as a significant issue in many countries, since clinically severe cases are more likely to be recognized and tested (17). Thus, our model was calibrated to match the trajectories of COVID-19-related deaths, hospitalized cases, and ICU bed use, as they are more reliable compared to data derived from diagnosed cases. Specifically, transmission rate, proportion of patients who would need hospital care or ICU, and the effect of NPI were varied until the model reproduced the observed trajectories. The end date was set to be May 8^th^, as this day was the end of the week in which lockdown restrictions were eased.

For each scenario, 1000 runs were performed. The 2.5 and 97.5 percentiles were also shown to include the appropriate uncertainty (stochastic variability). To measure the accuracy of the model’s prediction against the observed data, the least square method was used (i.e., minimize the square distance between observed and modeled data). Further details regarding the calibration procedure are available in the supplementary material.

### Examined scenarios

A ‘status quo’ scenario was used to generate predictions regarding the observed course of the outbreak. Additionally, two more scenarios were considered. Specifically, we implemented a hypothetical counterfactual scenario, wherein no interventions were implemented (neither social distancing measures nor increase in ICU bed capacity), in order to estimate how the outbreak would have unfolded if no interventions had taken place and a scenario wherein only the number of ICU beds had increased (ICU-only scenario).

## Results

### Model fit

Estimates regarding model’s fit are presented in the supplementary document. Model’s status quo scenario accurately captures the observed trends in hospitalized cases, ICU bed use, and COVID-19-related deaths (Supplementary, page 6, Figure S2-S4).

### The course of the first wave in Cyprus (March 1^st^- May 8^th^, 2020)

According to the model, the basic reproduction number (R0) was estimated at 2.66. The total number of COVID-19 cases in the Republic of Cyprus by May 8^th^ was 2690 (95% Credible Intervals (CrI): 1300, 4700) (Figure 1a). Using as denominator the number of total infections obtained from the model, the infection attack ratio of the first epidemic wave was 0.31% (95% CrI: 0.15%, 0.54%). Additionally, the case ascertainment ratio and the infection fatality ratio of the first epidemic wave were 33.2% (95% CrI: 19.7%, 68.7%) and 0.71% (95% CrI: 0.44%, 1.61%), respectively.

**Figure 1:**
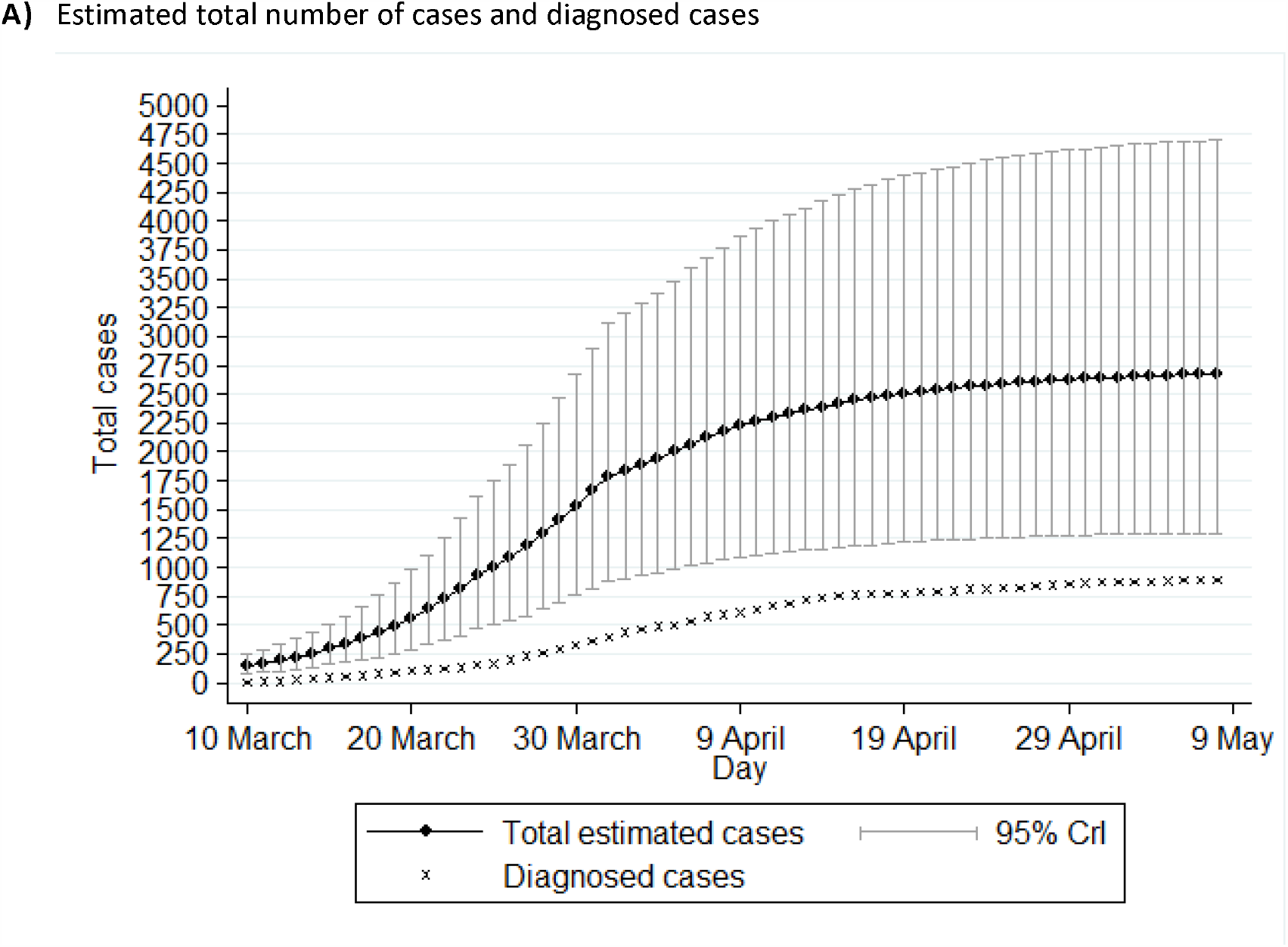
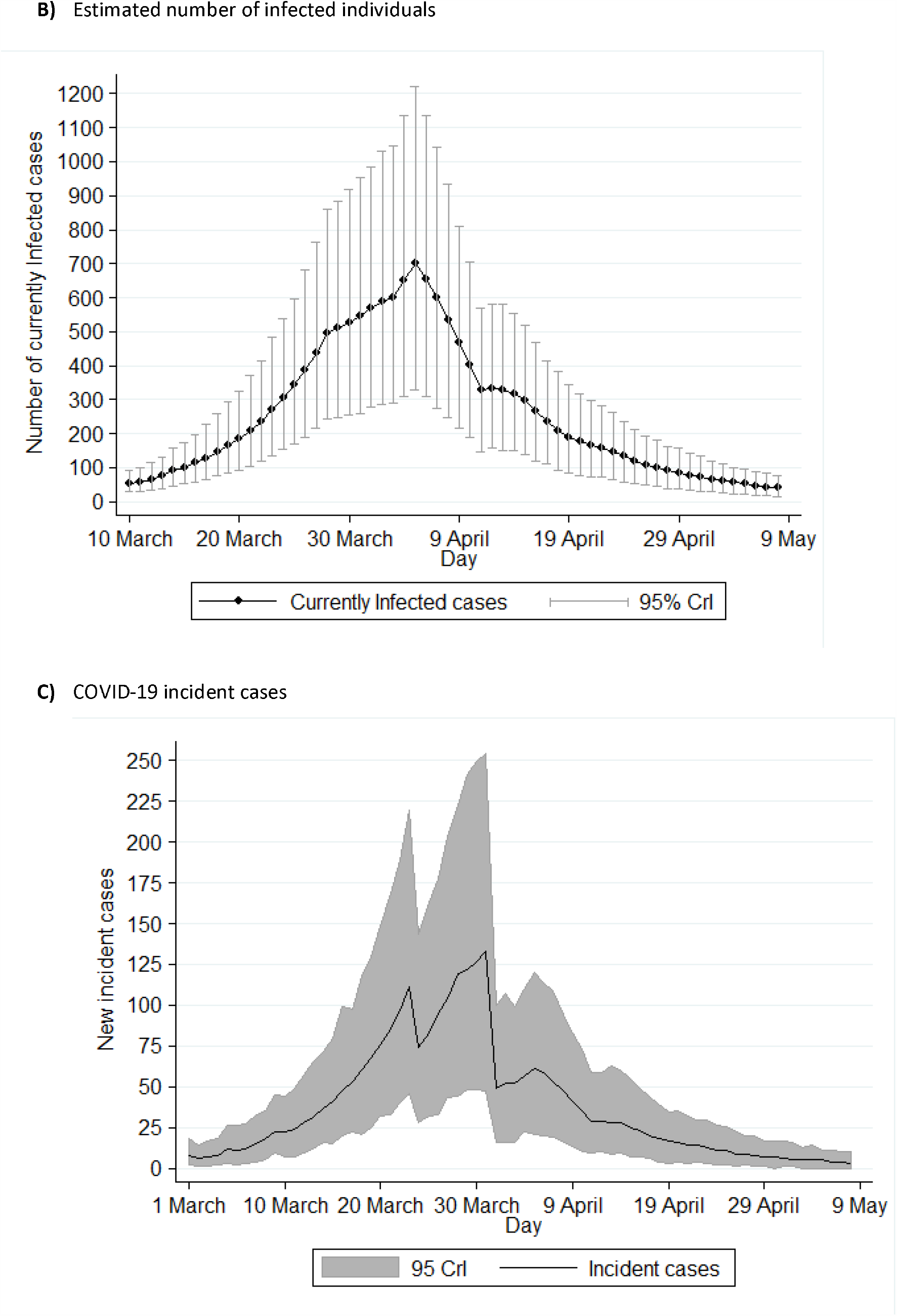
Model’s estimation regarding the first wave of COVID-19 in the Republic of Cyprus (1 March-8 May 2020): (A) Estimated total number of cases and diagnosed cases, (B) Estimated number of infected people; (C) COVID-19 incident cases. The error bars show the 95% credible intervals (95% CrI) for the model projections.

At the end of the study prediction dates (i.e., May 8^th^), it is estimated that there were 3 new infections per day (95% CrI: 0, 7) and 39 infectious cases (95% CrI: 15, 70). The model estimated that the number of new infections and the number of infected individuals peaked on March 31st and April 5th, respectively (Figure 1b and 1c).

### Counterfactual scenario

The model predicted that, under the counterfactual scenario, the total number of infected would be 86,600 (95% CrI: 46,700, 122,100) (Figure 2c). Additionally, the pressure on the healthcare system would have been substantially higher without NPI. More specifically, if the Republic of Cyprus had not taken any measure, the need for ICU beds would have exceeded the healthcare system capacity by April 14^th^ (Supplementary, page 8, Figure S6).

**Figure 2:**
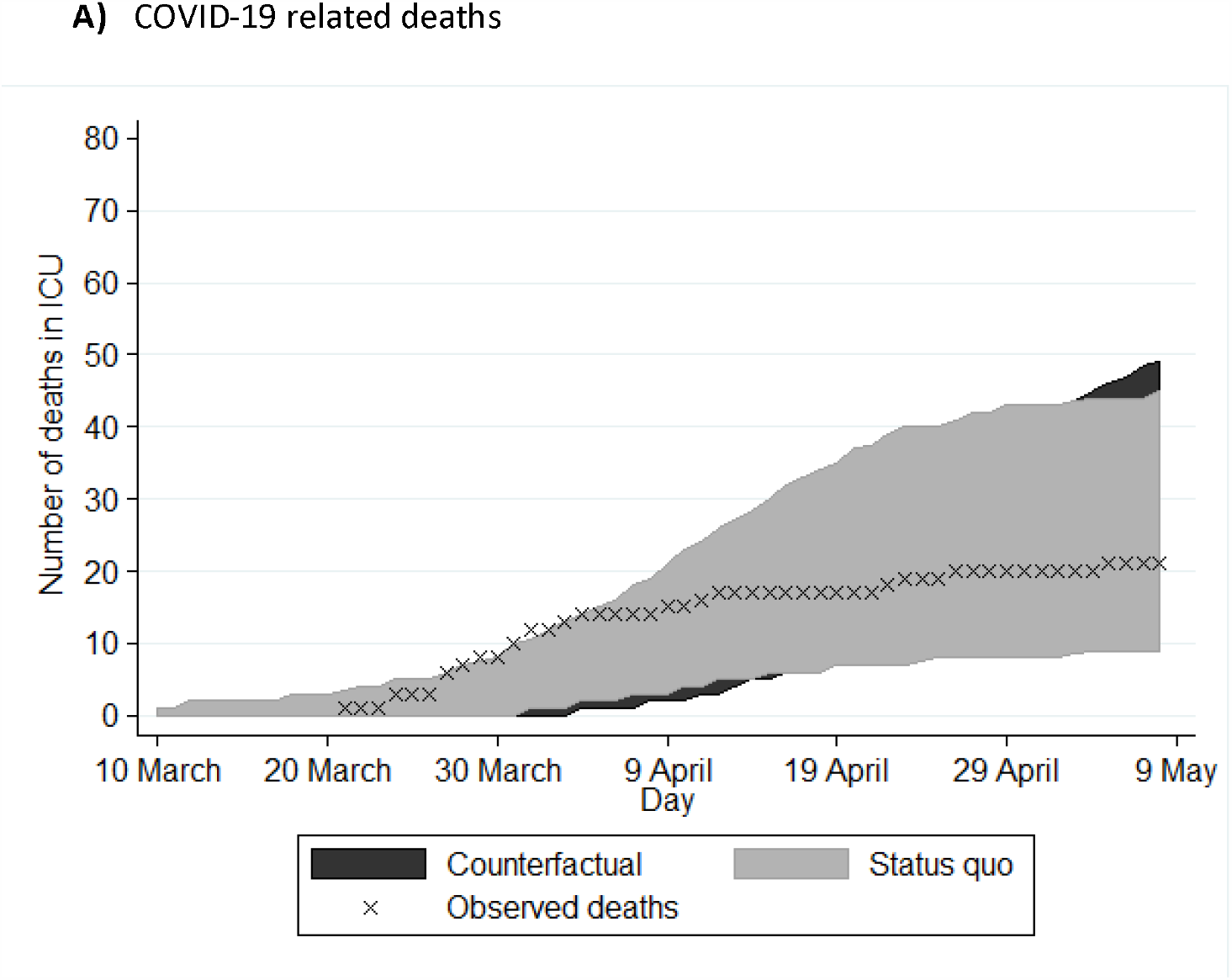
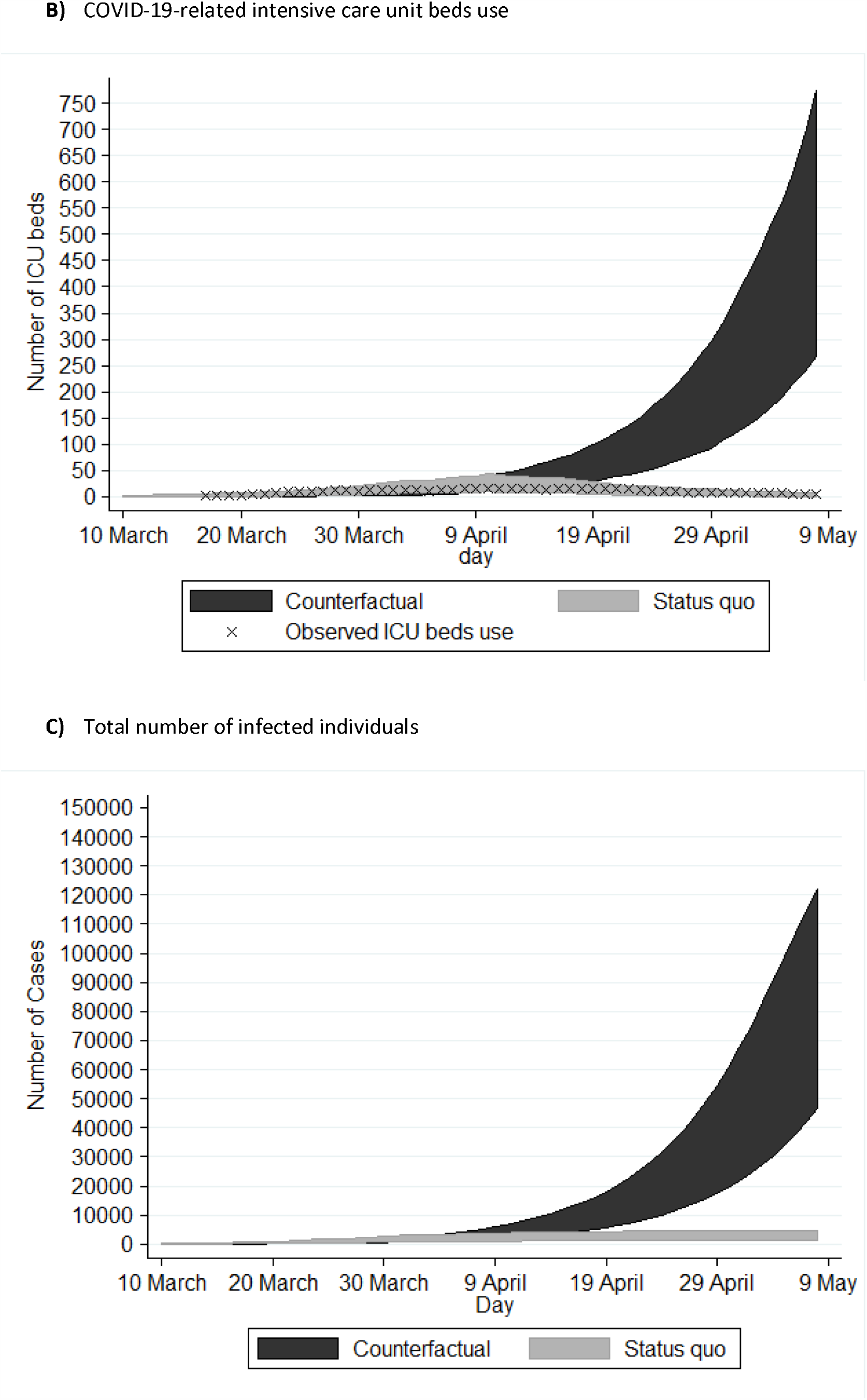
Projections of future COVID-19 cases and complications under status quo and counterfactual scenarios. For comparison, x indicates the observed trends under the status quo scenario: (A) Cumulative COVID-19 related deaths; (B) Daily COVID-19-related intensive care unit (ICU) beds use; and (C) Total number of infected individuals

Under the counterfactual scenario, the projected COVID-19-related number of deaths in ICUs would be 35 (95% CrI: 23, 49) by May 8^th^ (Figure 2a). Taking into account that additional deaths could occur due to non-availability of ICU beds, COVID-19-related mortality would be significantly higher. Specifically, assuming that 95% of those in need of an ICU bed would die in the absence of available ICU beds (18), the additional deaths by May 8^th^ would be 680 (95% CrI: 320, 1180) (Figure 2b). Thus, the expected number of deaths without any interventions (neither social distancing measures nor increase in ICU bed capacity) would be 715 (95% CrI: 339, 1235) by May 8^th^

### ICU-only scenario

If only ICU beds had increased from 27 to 50, the healthcare system would have lasted only 5 days longer from being overwhelmed, compared to the counterfactual scenario (Figure S7).

Under this scenario, the computed COVID-19-related number of deaths would be 58 (95% CrI: 40, 75) by May 8^th^ (Figure S7a) and the estimated deaths due to the non-availability of an ICU bed would be 615 (95% CrI: 270, 1080) (Figure S7b). Thus, if only ICU bed capacity had improved, the expected number of deaths would be 673 (95% CrI: 310, 1160) by May 8^th^.

## Discussion

Mathematical models can help understand SARS-CoV-2 transmission and assess the efficacy of measures to mitigate viral spread. Our model highlights that non-pharmaceutical interventions (both social distancing measures and increase in ICU bed capacity) were highly successful, as they kept the number of COVID-19-related deaths at low levels and the need for ICU beds within the capacity of the local health-care system.

IFR is a critical indicator for the evaluation of the consequences of the COVID-19 pandemic. A recent modeling study on different settings estimated an IFR ranging from 0.5 to 1.4% (19). Compared to those estimations, Cyprus experienced a low IFR (central estimation: 0.71%). Possible explanations for the country’s low IFR include: a) timely public health interventions, since NPI launched before the first death, b) opportunity to improve the preparedness of the country’s health-care system, since Cyprus was not among the first affected countries, and c) the geographic position of Cyprus, which is a peripheral country of the European Union and thus less vulnerable to the effect of external seeds.

According to our estimates, only a relatively small proportion of people in the Republic of Cyprus have been infected during the first epidemic wave (infection attack ratio: 0.31% (95% CrI: 0.15%, 0.54%), which is among the lowest in Europe (3). This means that Cyprus is far from achieving collective immunity (the collective immunity threshold for Cyprus was estimated at 1-(1/R0)=1-(1/2.66)=62.4%). Under the counterfactual scenario, which resembles a strategy based on herd immunity from natural infections, 86,600 individuals (95% CrI: 46,700, 122,100) would have been infected, which corresponds to a prevalence of 9.9% (95% CrI: 5.3%, 13.9%), still well below the target of 62.4%. Therefore, collective immunity could only be achieved in Cyprus through a combination of vaccination and infection, although the duration of immunity following infection remains unknown (20).

Globally, COVID-19 true infections are more than the laboratory-confirmed cases (3, 17). However, case ascertainment in the Republic of Cyprus during the first epidemic wave was relatively high (33.2%). This means that, during the first epidemic wave, each individual diagnosed with COVID-19 in Cyprus corresponded to about two undiagnosed cases. The corresponding ratios for Germany, Spain, Sweden, and Greece were 23.5%, 9.8%, 6.1%, and 24%, respectively (3, 7, 21). It is important that the efficacy of the testing strategy to control COVID-19 is strongly dependent on the efficiency of backward contact tracing and adherence of the infected individuals to isolation.

According to our model estimates, the implemented public health measures averted 715 (95% CrI: 339, 1235) deaths. Comparing the different interventions, social distancing measures were more important to flattening the epidemic peak and reducing the pressure on the health care system than the increase of the ICU bed capacity only. Consistent with previous work (7, 9), our results highlighted that any intervention to boost only ICU bed capacity would not have been an effective healthcare policy, as the demand for ICU beds would rapidly outrun availability. Figure 2b displays that after April 10^th^, the demand for ICU beds would have increased exponentially. However, despite the efficacy of NPI, it is important to take into consideration their indirect negative effects, including reductions in economic activities, mental issues due to isolation, and difficulties in accessing health care for chronic and other diseases (22). Thus, any social distancing measures should be implemented judiciously and only when the benefits outweigh the societal harms (22).

### Strengths and Limitations

This paper is useful as it provides theoretical support that fast and accurate interventions in the Republic of Cyprus during the first COVID-19 wave prevented the overload of the healthcare system. As with any modelling study, there are also limitations. First, the model ignores the impact of social networks in the population and assumes that it is randomly mixed. Second, an important assumption is that all deaths due to SARS-CoV-2 infection have been identified and reported (e.g., no deaths under the status quo scenario occurred before admission to the health-care facility). Third, we assumed that post-infection immunity completely protects again reinfection over the duration of simulations. Fourth, we assumed that all the deaths occurred in the ICU. Notwithstanding, the effect of this assumption on our projections is likely to be marginal, since the simple-bed mortality is relatively low. Finally, in the counterfactual scenario, public health interventions were removed, while assuming that everything else remained exactly as in the status quo scenario, i.e., there would be no changes in the duration that a patient stays in an ICU bed or in hospital.

## Conclusions

Non-pharmaceutical interventions, including increased testing rates and active contact tracing, limited the burden of the first wave of COVID-19 in the Republic of Cyprus and prevented the health-care system from becoming overwhelmed. Furthermore, our results highlight that collective (“herd”) immunity could not have been achieved in the Republic of Cyprus, even if social distancing measures were not taken. The findings of our study could be used as a guide in the confrontation of next waves of the COVID-19 in the Republic of Cyprus or other similar settings.

## Data Availability

All the data are in the manuscript

## Declarations

### Authors’ contributions

IG and GN conceived the study; IG performed the modelling and drafted the manuscript; GN coordinated the study; All authors provided essential inputs and contributed extensively to writing the manuscript; All authors contributed to model interpretation and approved the final version.

## References

1. World Health Organization. Emergency Committee regarding the outbreak of novel coronavirus (2019-nCoV). Geneva: WHO: 2020.

2. Li Q, Guan X, Wu P, Wang X, Zhou L, Tong Y, et al. Early Transmission Dynamics in Wuhan, China, of Novel Coronavirus-Infected Pneumonia. The New England journal of medicine. 2020;382(13):1199–207.

3. Flaxman S, Mishra S, Gandy A, Unwin HJT, Mellan TA, Coupland H, et al. Estimating the effects of non-pharmaceutical interventions on COVID-19 in Europe. Nature. 2020.

4. Epidemiological Surveillance Unit of the Ministry of Health C. National Situation Report. Coronavirus Disease 2019 (COVID-19) 2020 [updated 25/09/2020; cited 2020 14/10]. Available from: https://www.pio.gov.cy/coronavirus/pdf/ep1205ien.pdf.

5. Quattrocchi A, Mamais I, Tsioutis C, Christaki E, Constantinou C, Koliou M, et al. Extensive Testing and Public Health Interventions for the Control of COVID-19 in the Republic of Cyprus between March and May 2020. Journal of clinical medicine. 2020;9(11).

6. Prem K, Liu Y, Russell TW, Kucharski AJ, Eggo RM, Davies N, et al. The effect of control strategies to reduce social mixing on outcomes of the COVID-19 epidemic in Wuhan, China: a modelling study. The Lancet Public health. 2020.

7. Gountas I, Hillas G, Souliotis K. Act early, save lives: managing COVID-19 in Greece. Public health. 2020;187:136–9.

8. Kucharski AJ, Russell TW, Diamond C, Liu Y, Edmunds J, Funk S, et al. Early dynamics of transmission and control of COVID-19: a mathematical modelling study. The Lancet Infectious diseases. 2020;20(5):553–8.

9. N. M. Ferguson DL, G. Nedjati-Gilani, N. Imai, K. Ainslie, M. Baguelin, S. Bhatia, A. Boonyasiri, Z. Cucunubá, G. Cuomo-Dannenburg, A. Dighe, H. Fu, K. Gaythorpe, H. Thompson, R. Verity, E. Volz, H. Wang, Y. Wang, P. G. Walker, C. Walters, P. Winskill, C. Whittaker, C. A. Donnelly, S. Riley, A. C. Ghani. Impact of non-pharmaceutical interventions (NPIs) to reduce COVID-19 mortality and healthcare demand (Imperial College COVID-19 Response Team, 2020). 2020.

10. Agapiou S. AA, Baxevani A., Christofides T., Constantinou E., Hadjigeorgiou G., Nicolaides C., Nikolopoulos G., Fokianos K. Modeling of Covid-19 Pandemic in Cyprus. arXivLabs. 2020.

11. He X, Lau EHY, Wu P, Deng X, Wang J, Hao X, et al. Temporal dynamics in viral shedding and transmissibility of COVID-19. Nature medicine. 2020;26(5):672–5.

12. ECDC,,. Transmission of COVID-19 2020 [cited 2020 14/11]. Available from: https://www.ecdc.europa.eu/en/covid-19/latest-evidence/transmission.

13. Roman W, Wolfgang G, Michael S, Sabine Z, Marcel M, Daniela N, et al. Clinical presentation and virological assessment of hospitalized cases of coronavirus disease 2019 in a travel-associated transmission cluster. medRxiv. 2020;Published online March 5.

14. Armstrong RA, Kane AD, Cook TM. Outcomes from intensive care in patients with COVID-19: a systematic review and meta-analysis of observational studies. Anaesthesia. 2020;75(10):1340–9.

15. Lauer SA, Grantz KH, Bi Q, Jones FK, Zheng Q, Meredith HR, et al. The Incubation Period of Coronavirus Disease 2019 (COVID-19) From Publicly Reported Confirmed Cases: Estimation and Application. Annals of internal medicine. 2020;172(9):577–82.

16. Lu FS, Nguyen AT, Link NB, Lipsitch M, Santillana M. Estimating the Early Outbreak Cumulative Incidence of COVID-19 in the United States: Three Complementary Approaches. medRxiv : the preprint server for health sciences. 2020.

17. Lavezzo E, Franchin E, Ciavarella C, Cuomo-Dannenburg G, Barzon L, Del Vecchio C, et al. Suppression of a SARS-CoV-2 outbreak in the Italian municipality of Vo’. Nature. 2020;584(7821):425–9.

18. European Centre for Disease Prevention and Control. Baseline projections of COVID- 19 in the EU/EEA and the UK: update 2020 17 September 2020 Report No.

19. Hauser A, Counotte MJ, Margossian CC, Konstantinoudis G, Low N, Althaus CL, et al. Estimation of SARS-CoV-2 mortality during the early stages of an epidemic: A modeling study in Hubei, China, and six regions in Europe. PLoS medicine. 2020;17(7):e1003189.

20. Omer SB, Yildirim I, Forman HP. Herd Immunity and Implications for SARS-CoV-2 Control. Jama. 2020.

21. worldometers.info. COVID-19 CORONAVIRUS PANDEMIC 2020 [cited 2020 15/10]. Available from: https://www.worldometers.info/coronavirus/.

22. Lytras T, Tsiodras S. Lockdowns and the COVID-19 pandemic: What is the endgame? Scandinavian journal of public health. 2020:1403494820961293.

