## Supplementary Information for "Effect of public health interventions during the first epidemic wave of COVID-19 in Cyprus: a modelling study"

### **Schematic outline of the mathematical model for COVID-19 transmission**

**Figure S1**: Schematic outline of the mathematical model for COVID-19 transmission and progression. Individuals in the population begin as susceptible to infection with SARS-CoV-2. Once infected, and after a latent period of infection, individuals will either remain asymptomatic/develop mild symptoms, or develop severe symptoms requiring hospitalization or become critically ill requiring hospitalization and admission to intensive care unit (ICU). Hospitalized cases will recover. Cases in the ICU either die, D, or recover, R after spending a period of recovery time in a hospital bed. Individuals who are asymptomatic or experience mild symptoms will recover after a short duration moving into the recovered compartment, R.

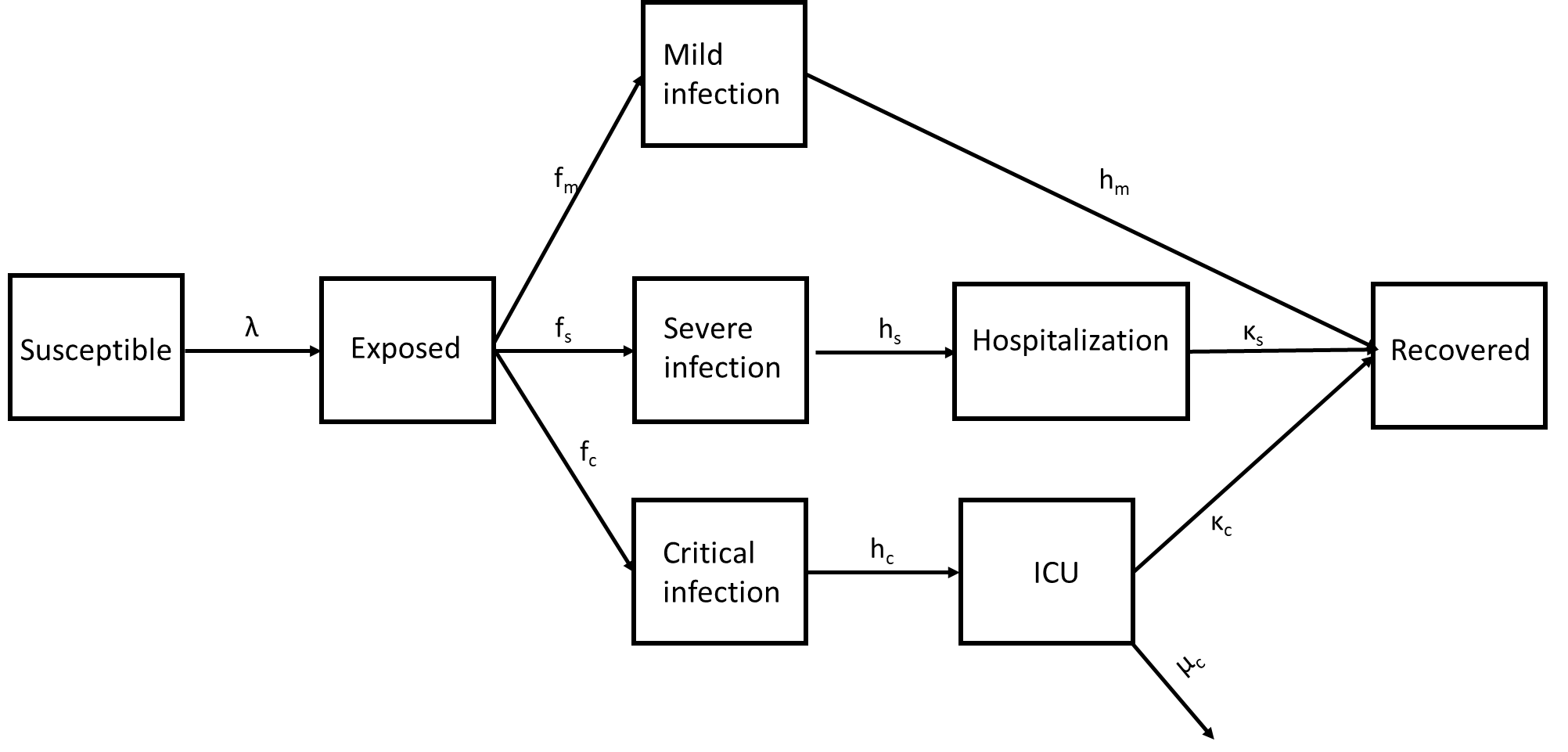

### **Description of a model through differential equations**

$$\frac{dS}{dt}=-(z_{i}*\lambda)*I_{Infectious\_stage}*(\frac{S}{N})$$

$$\frac{dE}{dt}=(z_{i}*\lambda)*I_{Infectious_{stage}}*\left( \frac{S}{N} \right)-\left( f_{m}+f_{s}+f_{c} \right)*E*\delta$$

$$\frac{d(Mild\_Infection)}{dt}=f_{m}*E*\delta-h_{m}*Mild\_Infection$$

$$\frac{d(Severe\_Infection)}{dt}=f_{s}*E*\delta-h_{s}*Severe\_Infection$$

$$\frac{d(Critical\_Infection)}{dt}=f_{c}*E*\delta-h_{c}*Critical\_Infection$$

$$\frac{d(Hospitalization)}{dt}=h_{s}*Severe\_Infection- {(k}_{s}+\mu_{s})*Hospitalization$$

$$\frac{d(ICU)}{dt}=h_{c}*Critical\_Infection- {(k}_{c}+\mu_{c})*ICU$$

$$\frac{d(R)}{dt}=h_{m}*Mild_{Infection}+k_{s}*Hospitalization+k_{c}*ICU$$

$$\frac{d(Death)}{dt}=\mu_{s}*Hospitalization+\mu_{c}*ICU$$

### **Explanation of symbols**

$I_{Infectious\_stage}$: Infectious population

λ: Force of Infection

$z_{i}$: Relative reduction of force of infection due to social distancing measures

1/δ: Latent period

$f_{m}$: Proportion of infections that will remain asymptomatic or develop mild symptoms

$f_{s}$: Proportion of infections that will progress to severe disease

$f_{c}$: Proportion of infections that will progress to critical illness

1/h_m_: Duration of mild infection

1/hs: Duration of severe disease before hospitalization

1/hc: Duration of severe disease before ICU

1/k_S_: Duration of hospitalization with severe disease

1/K_c_: Duration of ICU and hospitalization with critical illness

μ_s_: Disease mortality during hospitalization

μ_c_: Disease mortality in ICU

### **Model’s type**

In our analysis, a discrete-time, stochastic, individual-based model (IBM) was used. IBM simulate the patients’ trajectories at an individual level. It is important that those models possess some inherent randomness due to their methodology. The way that the model examines if a pseudo-individual would change state (e.g., from susceptible to infected) is through the draw of random numbers. More specifically, the model estimates the probability of moving from one stage to the next (e.g., from susceptible to infected). Then, for each pseudo-individual, a random number from a Uniform (0,1) distribution is drawn. If the resulted random number (e.g., 0.3) is smaller than the estimated probability of changing stage (e.g. 0.4), this pseudo-individual changes stage and vice versa. For example, regarding the transmission from susceptible to infected, if the risk of infection is 20%, then all the individuals with drawn random numbers lying in the range of (0-0.2) are assumed to become infected.

As the outcome of each run depends on chance, every simulation leads to slightly different results (Figure S2). Uncertainty comes from a single set of parameters but across multiple simulations with randomness included. For that, results over all simulations are pooled and the median along the 95% range are normally presented (stochastic variability). For more details regarding the IBM models one could look at (1).

**Figure S2**: Model predictions of ICU bed use under status quo scenario for the first 25 simulations (different colors) of model. The solid black line shows the median estimation.

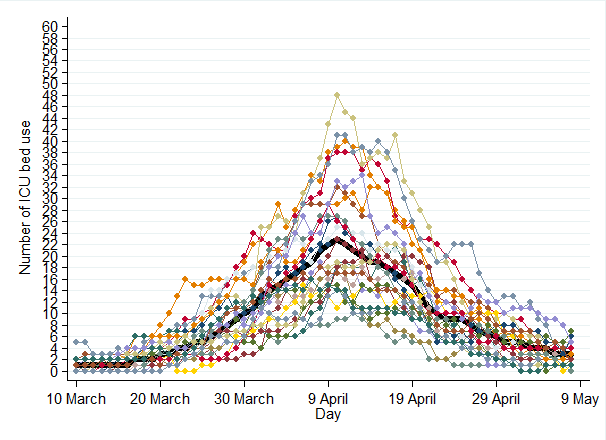

### **Goodness-of-fit metric (GoF)**

A GoF metric serves as the objective function in an optimization procedure, measuring the accuracy of the model’s predictions against the targets. The least square method was used to measure the accuracy.

GoF=∑(proj-obs)^2^

Smaller values of the GoF metric indicate a better fit to the observed data.

### **Model calibration to epidemiological and clinical data**

Fitting a model to empirical data (calibration procedure) improves the confidence that the simulation outputs are realistic and accurate. A back-calculation methodology was used to estimate the course of the epidemic from observed deaths, hospitalized cases, and ICU cases. A three-step repetitive process was applied to estimate the optimal combination of the model’s parameters. The calibration procedure of our model is described below.

**Estimation of baseline parameters**

The first social distancing measure in the Republic of Cyprus was launched on March 10^th^, 2020 (schools and universities closure). Given that there is a **10 days-time-lag** between the introduction of control measures and their subsequent impact on morbidity and mortality, the number of hospitalized cases, the number of patients in ICU beds, and the COVID-19-related deaths by March 19^th^ were used in combination to estimate our baseline parameters: (a) basic reproduction number, b) proportion of the population that would need hospitalization, and c) proportion of the population that would need ICU bed if infected.

Initially, a combination that fits well the observed COVID-19 epidemiological data by March 19^th^ was manually found. Specifically, the initial values for the transmission rate (b), the proportion of the population in need of hospitalization, and the proportion of people in need of ICU beds were 0.39, 11%, and 2.2%, respectively. Then, our initial values were updated through an iterative process. Specifically, we examined all the possible combinations ranged between the ±20% of the initial values (e.g., possible solutions of the transmission rate ranged between 0.31 and 0.47; the proportion of the population in need for hospitalization from 9 to 14%, and the proportion of the population in ICU from 2 to 3%). In total, 12800 different parameter combinations were assessed (all the iteration steps for b, hospitalization need, and need of ICU beds were 0.5, 0.25, and 0.25, respectively). Per simulated scenario, one hundred model runs were performed. To determine which set of parameters provides the optimal fit to the epidemiological data, the least square method was used (minimize the square distance between observed and modeled data). The final the values for the transmission rate (b), the proportion of the population in need of hospitalization and the proportion of people in need of ICU beds were 0.41, 11.5% and 2.5%, respectively.

Our model showed that in the Republic of Cyprus 14% (i.e., 11.5%+2.5%) of the patients would need to enter the healthcare system. This figure is similar with the observed proportions that were computed for the Chinese population that needed to enter the healthcare system (13.5%) (2-4).

**Effect of social distancing measures**

Finally, in order to consider the impact of social distancing measures in the model, the infection rate was multiplied by the parameter-vector **z** (z_i_ ranges between 0 and 1 and reflects the different steps in measures taken). We considered the effect of 4 social distancing measures (closure of school, restricted access to the country, closure of shops and restaurants, and total lockdown). Specifically, we adjusted the values of **z** to obtain the optimal fit to the observed ICU beds and COVID-related deaths **post** March 19^th^ (after March 19^th^, the effect of the measure could be observed in ICU beds and COVID-related deaths). To assess the optimal fit, the least square method was also used (minimize the square distance between observed and modeled data).

**Table S1**: Timeline of the implemented public health measures to mitigate the spread of SARS-CoV-2

| **Timeline** | **Measures** |
| --- | --- |
| March 10^th^ | School and university closures, public gathering (>75 persons) cancelled |
| March 15^th^ | Restricted access to the country, closure of entertainment areas, 1 person per 8 square meters in public services areas |
| March 24^th^ | Restriction in Construction sites, Closure of the majority of retail services |
| March 31^st^-May 3^rd^ | Incoming flights suspended but for repatriated citizens, compulsory 14-day quarantine for inbound travelers, inter-city travel and all social gatherings prohibited, inner-city movements restricted, night curfew, limited mobility permissions per person per day |

A back-calculation methodology to estimate the course of the epidemic from observed deaths was also used from the Imperial College Covid19 Response Team (5).

### **Model fit**

Figures S2-S4 show that the model’s status quo scenario accurately captures the overall trends in ICU bed use and COVID-19 related deaths.

**Figure S3**: ICU beds use under the status quo scenario. For comparison, asterisks indicate the observed trends

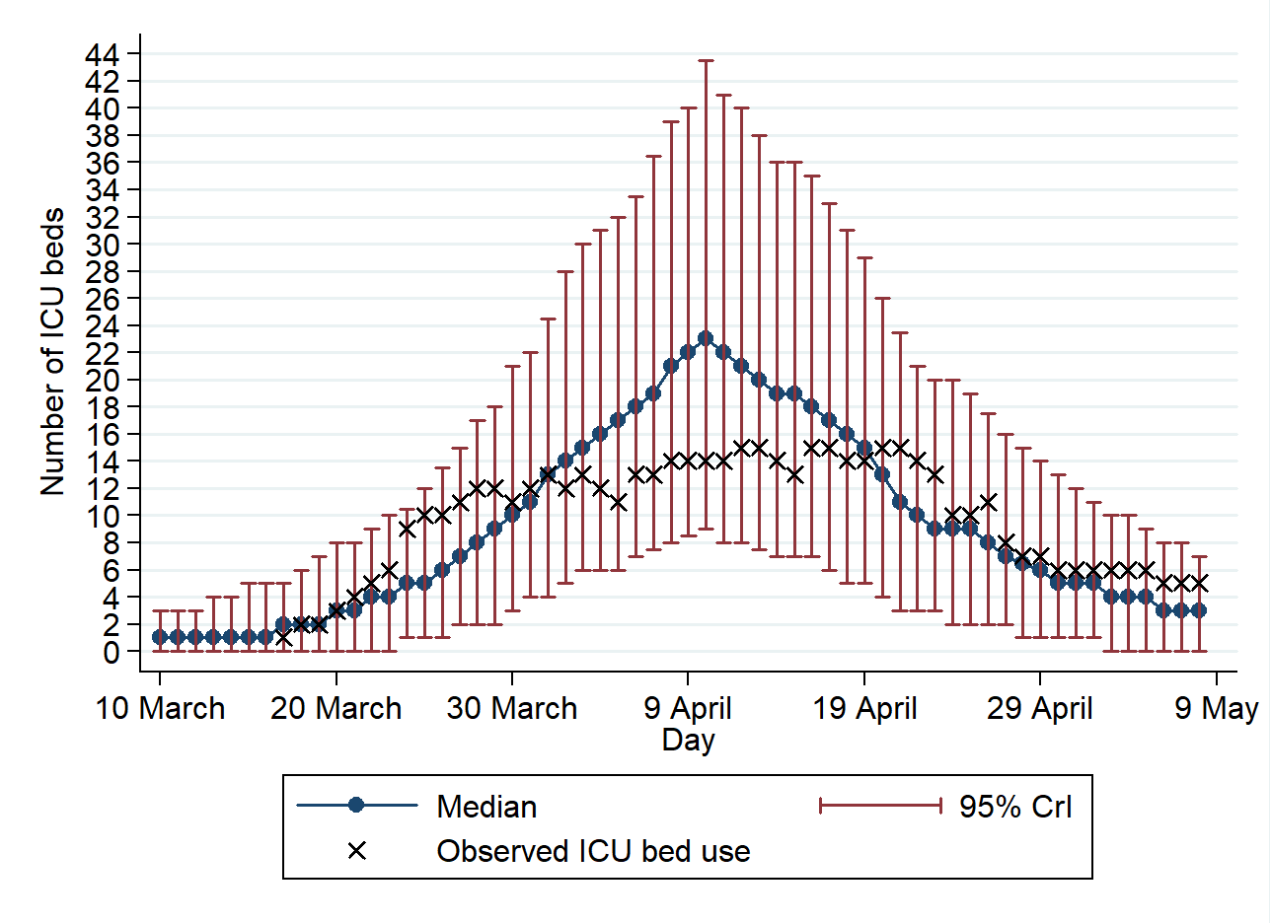

**Figure S4**: COVID-19 related deaths under the status quo scenario. For comparison, asterisks indicate the observed trends

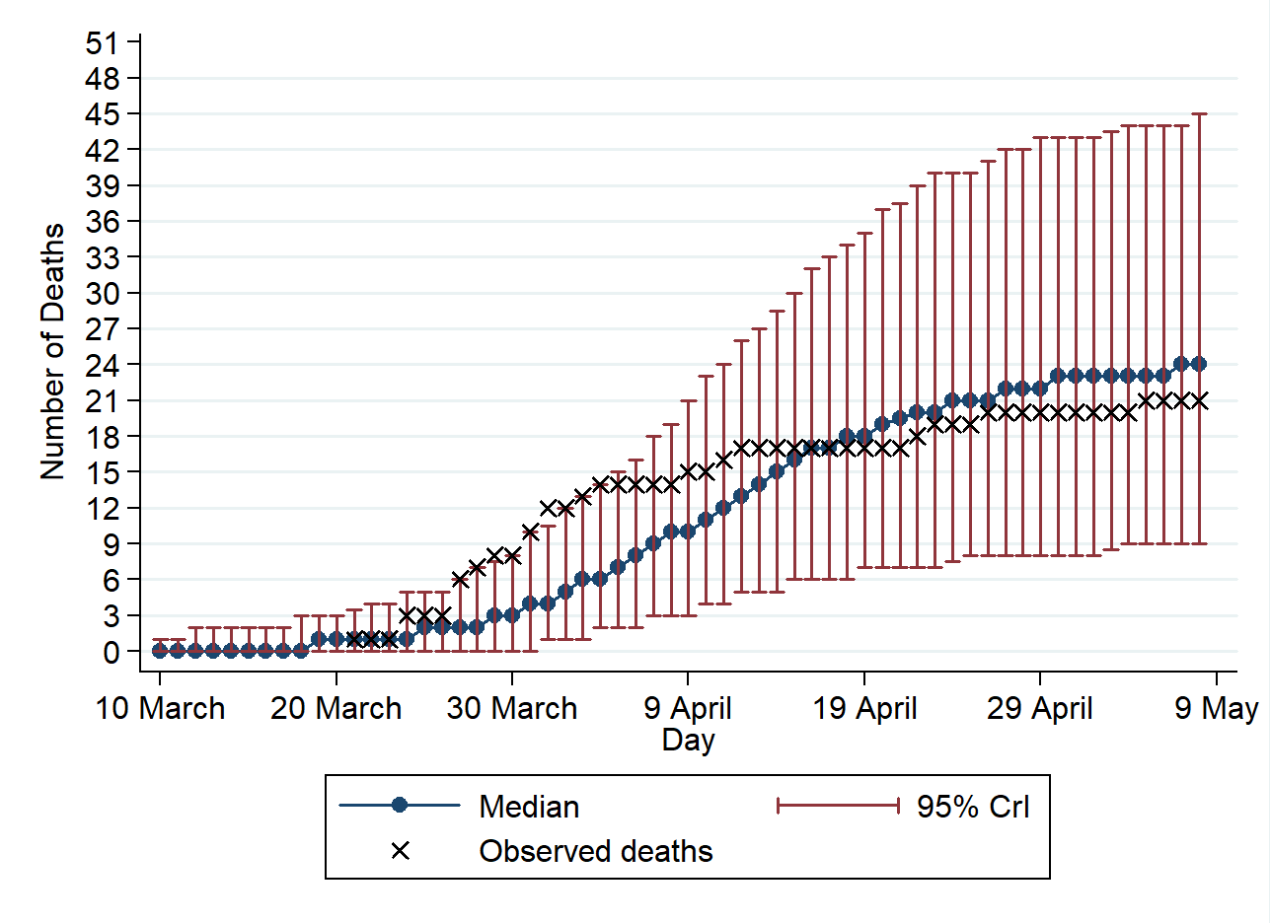

**Figure S5**: Hospitalized cases under the status quo scenario. For comparison, asterisks indicate the observed trends

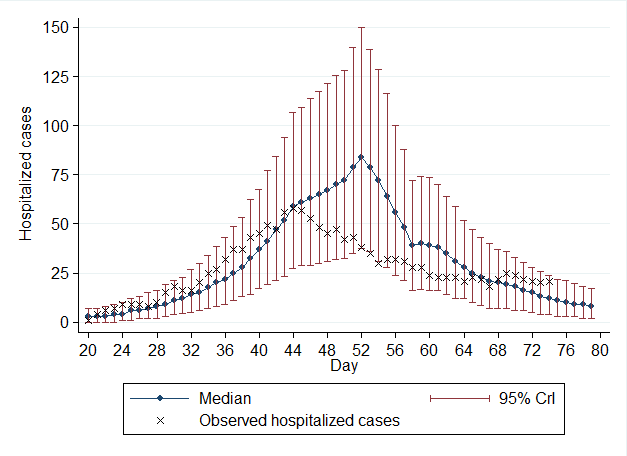

### **Estimated time for the healthcare system to become overwhelmed under counterfactual scenario**

Without non-pharmaceutical interventions, the strain on the healthcare system in the Republic of Cyprus, due to COVID-19, would be significantly higher. More specifically, without non-pharmaceutical interventions, the need for ICU beds would have surpassed the healthcare system capacity April 14^th^.

**Figure S6**: ICU beds use under the counterfactual scenario.

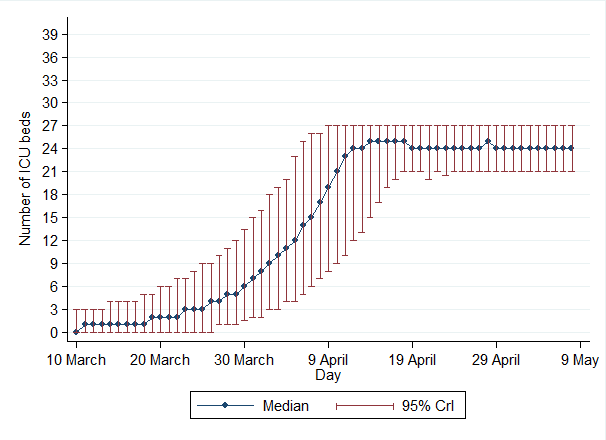

**What would have happened if only the ICU bed capacity had improved?**

In case the ICU bed capacity had increased, without any non-pharmaceutical measures, the need for ICU beds would have exceeded the healthcare system capacity by April 19^th^.

**Figure S7**: ICU beds use under the counterfactual scenario.

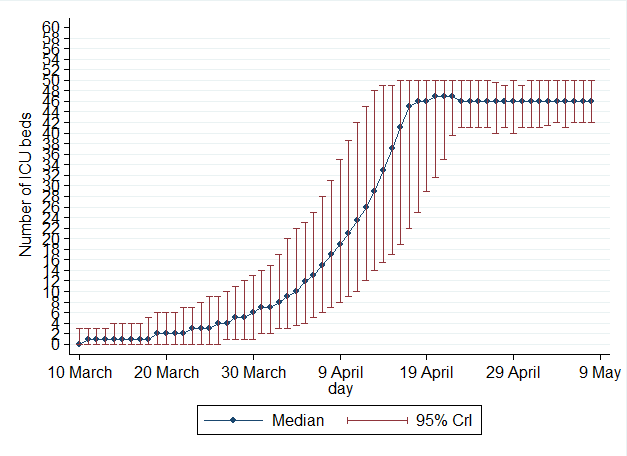

**Figure S8:** Projections of COVID-19 cases and complications under the status quo scenario and the scenario where only ICU bed capacity increased. For comparison, x indicates the observed trends under the status quo scenario. (A) Cumulative COVID-19-related deaths and (B) Daily COVID-19-related intensive care unit (ICU) beds use

A) **COVID-19 related deaths**

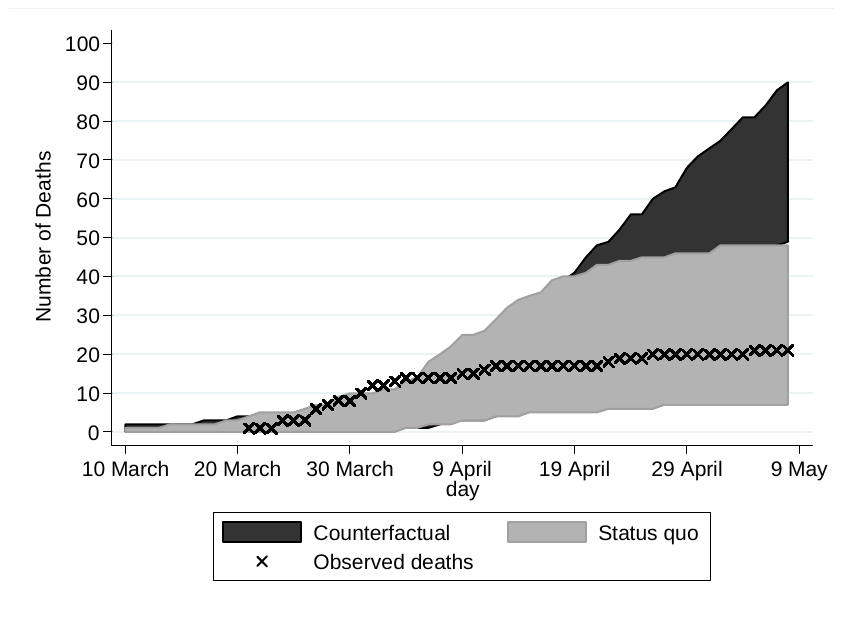

B) **COVID-19-related intensive care unit beds use**

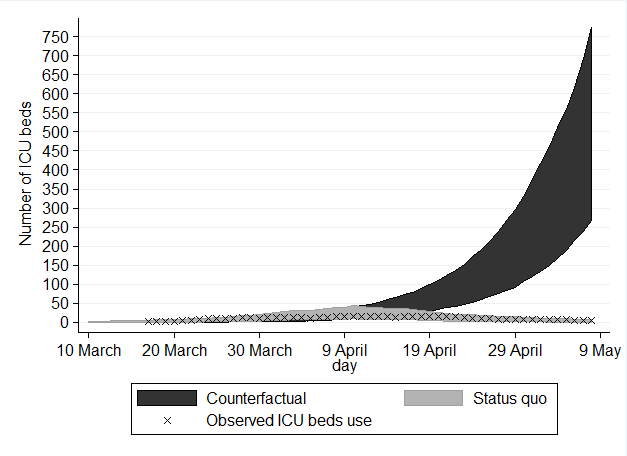

### **References**

1.	Vynnycky Emilia , Richard White An Introduction to Infectious Disease Modelling Illustrated Edition, Kindle Edition2010.

2.	Guan WJ, Ni ZY, Hu Y, Liang WH, Ou CQ, He JX, et al. Clinical Characteristics of Coronavirus Disease 2019 in China. The New England journal of medicine. 2020;382(18):1708-20.

3.	Huang C, Wang Y, Li X, Ren L, Zhao J, Hu Y, et al. Clinical features of patients infected with 2019 novel coronavirus in Wuhan, China. Lancet. 2020;395(10223):497-506.

4.	Ayoub HH, Chemaitelly H, Seedat S, Mumtaz GR, Makhoul M, Abu-Raddad LJ. Age could be driving variable SARS-CoV-2 epidemic trajectories worldwide. PloS one. 2020;15(8):e0237959.

5.	Flaxman S, Mishra S, Gandy A, Unwin HJT, Mellan TA, Coupland H, et al. Estimating the effects of non-pharmaceutical interventions on COVID-19 in Europe. Nature. 2020.
